# Prevalence, Determinants and Care Seeking Behavior for Anxiety and Depression in Nepalese Population: An Analysis of Nepal Demographic and Health Survey Data 2022

**DOI:** 10.1101/2023.07.22.23293031

**Authors:** Achyut Raj Pandey, Bikram Adhikari, Bihungum Bista, Bipul Lamichhane, Deepak Joshi, Saugat Pratap KC, Shreeman Sharma, Sushil Chandra Baral

## Abstract

**Objective:** To determine the prevalence and factors associated with anxiety and depression and the care seeking behaviour among Nepalese population.

**Methods:** We analyzed secondary data from nationally representative Nepal health demographic survey 2022. Depression and anxiety were assessed using Patient Health Questionnaire-9 (PHQ-9) and Generalized Anxiety Disorder Assessment (GAD-7) tools respectively. We performed weighted analysis to account complex survey design. We presented categorical variables as frequency, percent and 95% CI whereas numerical variables as median, interquartile range and 95% CI around median. We performed univariate and multivariable logistic regression to determine factors associated with anxiety and depression, and results were presented as crude odds ratio (COR), adjusted odds ratio (AOR) and their 95% CI.

**Results:** The prevalence of depression was 4.0% (95%CI: 3.5, 4.5) in both sexes, 5.4% (95%CI: 4.8, 6.1) among females and 1.7% (95%CI: 1.4, 2.3) among males. Similarly, the prevalence of anxiety was 17.7% (95%CI: 16.5, 18.9) in both sexes, 21.9% (95%CI: 20.4, 23.6) among females and 11.3% (95%CI: 10.0, 12.8) among males. Divorced or separated participants were found to have higher odds of developing anxiety (AOR=2.39, 95% CI: 1.73, 3.30) and depression (AOR=3.11, 95% CI: 1.81, 5.35). Among different ethnic groups, Janajati had lower odds of developing anxiety (AOR=0.77, 95% CI: 0.64, 0.91) and depression (AOR=0.67, 95% CI: 0.48, 0.92) compared to Brahmin/Chhetri. Regarding disability, participants with some difficulty had higher odds of developing anxiety (AOR=1.82, 95% CI: 1.57, 2.10) and depression (AOR=1.96, 95% CI: 1.52, 2.51), and those with a lot of difficulty/can’t do at all had higher odds of anxiety (AOR: 2.10, 95% CI: 1.48, 2.97) and depression (AOR: 2.06, 95% CI: 1.07, 3.94) compared to those without any disability.

**Conclusion:** The prevalence of depression and anxiety were relatively higher among females compared to males. Marital status and disability status are positively associated with anxiety and depression whereas Janajati ethnicity and males were negatively associated with anxiety and depression. It is essential to develop interventions and policies targeting females and divorced individuals which can be helpful in reducing the burden of anxiety and depression in Nepal.

**Strengths and Limitations:**

1. *We analyzed data from large scale nationally representative survey that takes into consideration the recently federalized structure of the country*.
2. *Anxiety and depression have been assessed using PHQ-9 and GAD-7 tools that improves the comparability of findings with other studies*
3. *Weighed analysis was carried out to account complex survey design of the survey*
4. *The survey was during COVID-19 pandemic period which may have altered the prevalence of disease conditions to some extent*

## Introduction

In 2019, around 970 million people globally were estimated to be living with mental disorders, with approximately 82% of these cases being from Low- and Middle-Income countries (LMICs).^1^ On a global scale, one out of every eight people suffers from a mental disorder, with anxiety and depressive disorders being the most common.^2, 3^ There were 45.82 million cases of anxiety incident with estimated number of prevalent cases standing at 301.39 million in 2019. Anxiety disorders were responsible for approximately 28.68 million Disability-Adjusted Life Years (DALYs), with approximately 50% increase in absolute burden since 1990.^4^

Similarly, there were 280 million prevalent cases of depression with a prevalence rate of 3613.67 cases per 100,000 population, The number of prevalent cases increased by 63.17% since 1990. In 2019, there were 46.8 million DALYs from depression with approximately 61% increase from year 1990. ^3^ After back and neck pain, depression was second leading cause of years lived with disability (YLDs) in 2019 accounting approximately 5.6% of total YLDs while anxiety disorders stood at 6^th^ rank with approximately 3.4% of total YLDs worldwide.^1^

There were approximately 1.36 million prevalent cases of depression and 0.97 million prevalent cases of anxiety disorder in Nepal.^3^ The study estimates the prevalence of depressive disorder was 4.47% while anxiety disorder was 3.17% in 2019. Similarly, there were estimated 243,462 DALYs from depression and 91,927 DALYs from anxiety in 2019.^3^

Excessive fear and worry, along with behavioral disruptions, are characteristic of anxiety disorders. An array of anxiety disorders exist, encompassing conditions excessive worry, panic attacks, excessive fear and worry in social situations, extreme fear or anxiety regarding separation from emotionally attached individuals, and others.^2^ Individuals diagnosed with anxiety disorders can experience a frequently prolonged response when exposed to seemingly innocuous stimuli. This response is typically marked by sensations of tension, increased vigilance, activation of the sympathetic nervous system, subjective feelings of fear, and in certain circumstances, the onset of panic.^5^ Depression differs from normal mood swings and transient emotional responses. It is characterized by persistent feelings of sorrow, anger, or emptiness, as well as loss of interest or pleasure in activities that continue most days, for at least two weeks. Overwhelming guilt or poor self-worth, hopelessness, suicidal thoughts, sleep disturbances, changes in appetite or weight, and exhaustion are among other symptoms of depression.^2^

Anxiety is associated with increased disability and diminished health and wellbeing. Increased disability and diminished health and wellbeing are linked to anxiety.^6^ Anxiety has been found to be associated with multiple other health conditions like heart disease, depression, asthma and gastrointestinal problems.^7^ Globally, individuals with poor mental state are found to bear disproportionately higher burden of mortality compared to general population. ^1^ Calculation of mortality attributable to mental disorder including anxiety and depression is complex as mental disorders are rarely recorded as causes of deaths in death certificate. However, a report from World Health Organization reports that people with poor mental health die 10-20 years earlier than the general population. ^1^

Depression and anxiety are projected to cost the global economy $1 trillion each year, mostly owing to productivity losses. Despite the importance of economic activity in healing, persons with severe mental health issues are frequently excluded from the labour force.^8^ Mental health, of which major share is born by depression and anxiety are often less prioritized in research activities. Despite serious impact on health and wellbeing of individuals, mental health receives approximately 7% of global health research fundings. ^1, 9^ Approximately, 99% of mental health studies are funded by high income countries and only 5% of total mental health research finding go in LMICs^1, 9^ like Nepal. Although there are some studies anxiety and depression in Nepal, they are mostly confined in specific geographic area and among specific group of people like health care workers^10–13^, traffic police^14^, among patients with specific disease conditions^15–18^ specific geographic area.

There are limited studies carried out in a nationally representative sample of population to determine prevalence and factors associated with depression anxiety. In this study we aimed to determine prevalence and factors associated with depression and anxiety. In addition, we aimed to determine health seeking pattern among participants with anxiety and depression.

## Methods

### Study design

We analyzed data from nationally representative Nepal Health Demographic Survey, 2022 (NDHS 2022).

### Study setting

Nepal is a landlocked country located in Southeast Asia region with one federal, seven provincial and 753 local governments (6 metropolitan cities 11 sub-metropolitan cities, urban 275 municipalities, 460 rural municipalities). Nepal has three ecological region-Mountain, Hill and Terai. Based on Census 2021, the total population of Nepal was 29,164,578 with 911,027 (51.1 %) females and 14,253,551 (48.9 %) males. Nepal has the human development index of 0.602, inequality adjusted HDI of 0.449, planetary pressure adjusted HDI of 0.584 and ranks 143 in HDI among countries across the world.

## Sample and sampling

NDHS 2022 uses an updated sampling framework based on Housing and Population Census 2011. In the first stage of sampling, the seven provinces were stratified into rural and urban settings that together formed a total of 15 sampling stratum across seven provinces. Within each stratum, the sampling procedure included implicit stratification and proportionate allocation. The sampling frame was sorted inside each stratum based on administrative units, using a probability-proportional-to-size technique. A total of 476 primary sample units (PSUs) were chosen, with 248 from urban and 228 from rural settings. PSUs were chosen individually based on their size within each stratum. A household listing operation was carried out within each PSU and the resulting household list was considered as a sampling frame. Wards with a number of households more than 300 were further segmented and a segment was selected based on probability proportionate to size. From each cluster, a total of 30 households resulting in a total of 14,280 households, 7,440 were in urban areas and 6,840 from rural settings. A total of 14,845 women and 4,913 men were successfully interviewed. Detail sampling process is elaborated elsewhere.^19^

### Data collection

From January 5 to June 22, 2022, a total of 19 teams, each comprising a supervisor, one male interviewer, three female interviewers, and one biomarker specialist, conducted data collection for the NDHS 2022.

## Variables

### Dependent variables

#### Anxiety

NDHS 2022 used Generalized Anxiety Disorder Assessment (GAD-7) tool consisting of seven items to assess anxiety. Each item of GAD-7 was scored on a scale of 0 to 3 on a 4-point Likert scale (0 = not at all, 1 = several days, 2 = more than half the days and 3 = nearly every day). The scores from seven items were summed up to determine the total GAD-7 score. The total score of GAD-7 ranges from 0 to 21 (score of 0-5 is categorized as no anxiety, 6-14 as mild to moderate anxiety, 15-21 as “severe anxiety”). In this study, we considered participants have anxiety if GAD-7 score is >5.

#### Depression

NDHS 2022 used Patient Health Questionnaire-9 (PHQ-9) tool consisting of nine items to assess depression. Each item of PHQ-9 was scored on a scale of 0 to 3 in a 4-point Likert scale (0D=Dnot at all; 1D=Dseveral days; 2D=Dmore than a week; 3D=Dnearly every day). Scores from each item were summed-up to determine the total PHQ-9 score. The total score of PHQ-9 ranged from 0 to 27 (score of 0-5 is classified as “no depression”, scores of 5–9 is classified as mild depression; 10–14 as moderate depression; 15–19 as moderately severe depression; ≥ 20 as severe depression). In this study, we considered participants have depression if PHQ-9 score is >=10.

### Independent variables

The independent variables assessed in this study included age(<20 years, 20-34 years, 34-49 years), sex (male, female), marital status (unmarried, married or living together, divorced or separated or not living together), ethnicity (Brahmin/Chhetri, Dalit, Janajati, Madhesi, Other), religion (Hindu, Other), wealth quintile (poorest, poorer, middle, richer, richest), disability (no disability, some difficulty, a lot of difficulty or can’t do at all), education (no education, basic, secondary, higher), occupation (not working, agriculture, professional or technical manager or clerical, sales and service, skilled/unskilled labor, other), smoking (do not smoke, some day, everyday), alcohol(never drinker, no drink in past month, some drink in past month, everyday drink), ecological belt (mountain, hill, terai), place of residence (rural, urban), and province(Koshi, Madhesh, Bagmati, Gandaki, Lumbini, Karnali, Sudurpaschim).

### Statistical analysis

We used R version 4.2.0 and RStudio for data cleaning and statistical analysis. We performed weighted analysis to account complex survey design of NDHS 2022. We presented categorical variables as frequency, percent (%) and 95% confidence interval (CI) whereas numerical variables as mean and 95% CI. We used univariate and multivariable logistic regression to determine the association of depression and anxiety with independent variables. The results of the logistic regression were presented as crude odds ratio and adjusted odds ratio and their 95% CI. A p-value of <0.05 was considered statistically significant.

### Ethical approval

Ethical approval for the survey was obtained from Ethical Review Board of Nepal Health Research Council (Reference number: 678, Date: 30^th^ September 2021) and institutional review board of ICF international (Reference number: 180657.0.001.NP.DHS.01, Date: 28^th^ April 2022).

## Results

Slightly more than two-third (69.2%) of participants were from urban settings. Terai belt contributed the highest proportion of participants (55.1%), followed by Hill (39.5%) while 5.4% participants were from mountain region. Among the seven provinces, 22% of participants were from Bagmati, 20.4% from Madhesh, 17.6% from Lumbini, 17.2% from Koshi, 8.9% from Gandaki, 8.1% from Sudurpaschim, and 5.9% from Karnali. Janajati accounted for the highest proportion of participants (37.4%), followed by Brahmin/Chhetri (26.6%) and Madheshi (16.9%). The majority of participants (82.6%) identified as Hindu. The median age of participants was 29 years, with 46.8% falling within the 20-34 age group. In terms of marital status, 70.1% of participants were married and living together, 27.6% were unmarried, and the remaining 2.3% were divorced or separated. Approximately 19% of participants had no education, and 21.9% were not currently employed. Around 83.5% of participants were non-smokers, while 13.9% were lifetime abstainers from alcoholIn terms of disability, 17.9% of participants had some level of difficulty. Only 12.6% of participants had health insurance coverage.

**Table 1:** Characteristics of research participants (n=12,322)

| Characteristic | Overall |  | Female |  | Male |  |
| --- | --- | --- | --- | --- | --- | --- |
|  | n | % (95%CI) | n | % (95%CI) | n | % (95%CI) |
| **Place of residence** |  |  |  |  |  |  |
| Urban | 8,525 | 69.2 (67.6, 70.7) | 5,064 | 68.3 (66.8, 69.9) | 3,462 | 70.5 (68.6, 72.3) |
| Rural | 3,796 | 30.8 (29.3, 32.4) | 2,345 | 31.7 (30.1, 33.2) | 1,451 | 29.5 (27.7, 31.4) |
| **Ecological belt** |  |  |  |  |  |  |
| Mountain | 663 | 5.4 (3.75, 7.67) | 408 | 5.5 (3.86, 7.82) | 255 | 5.2 (3.55, 7.53) |
| Hill | 4,869 | 39.5 (35.6, 43.5) | 2,896 | 39.1 (35.2, 43.1) | 1,973 | 40.2 (36.0, 44.5) |
| Terai | 6,790 | 55.1 (51.2, 58.9) | 4,105 | 55.4 (51.5, 59.2) | 2,685 | 54.6 (50.4, 58.8) |
| **Province** |  |  |  |  |  |  |
| Koshi | 2,123 | 17.2 (15.9, 18.6) | 1,241 | 16.8 (15.4, 18.2) | 882 | 18 (16.4, 19.6) |
| Madhesh | 2,509 | 20.4 (19.1, 21.7) | 1,512 | 20.4 (19.1, 21.8) | 997 | 20.3 (18.7, 22.0) |
| Bagmati | 2,707 | 22 (20.2, 23.8) | 1,493 | 20.1 (18.5, 21.9) | 1,214 | 24.7 (22.3, 27.2) |
| Gandaki | 1,092 | 8.9 (7.83, 10.0) | 704 | 9.5 (8.36, 10.8) | 387 | 7.9 (6.88, 9.00) |
|  | n | % (95%CI) | n | % (95%CI) | n | % (95%CI) |
| Lumbini | 2,171 | 17.6 (16.3, 19.0) | 1,359 | 18.3 (17.0, 19.8) | 812 | 16.5 (15.0, 18.2) |
| Karnali | 724 | 5.9 (5.31, 6.50) | 458 | 6.2 (5.62, 6.80) | 266 | 5.4 (4.72, 6.21) |
| Sudurpashchim | 996 | 8.1 (7.37, 8.85) | 641 | 8.7 (7.96, 9.40) | 355 | 7.2 (6.32, 8.24) |
| **Ethnicity** |  |  |  |  |  |  |
| Brahmin/Chhetri | 3,281 | 26.6 (24.3, 29.1) | 2,049 | 27.7 (25.3, 30.1) | 1,232 | 25.1 (22.5, 27.8) |
| Dalit | 1,773 | 14.4 (12.7, 16.2) | 1,115 | 15.1 (13.3, 16.9) | 658 | 13.4 (11.6, 15.4) |
| Janajati | 4,604 | 37.4 (34.6, 40.2) | 2,734 | 36.9 (34.1, 39.8) | 1,869 | 38 (35.0, 41.2) |
| Madheshi | 2,066 | 16.8 (14.6, 19.2) | 1,149 | 15.5 (13.3, 18.0) | 917 | 18.7 (16.3, 21.3) |
| Others | 599 | 4.9 (3.48, 6.75) | 362 | 4.9 (3.44, 6.90) | 236 | 4.8 (3.41, 6.76) |
| **Religion** |  |  |  |  |  |  |
| Hindu | 10,174 | 82.6 (80.4, 84.6) | 6,149 | 83 (80.8, 85.0) | 4,025 | 81.9 (79.5, 84.1) |
| Other | 2,147 | 17.4 (15.4, 19.6) | 1,259 | 17 (15.0, 19.2) | 888 | 18.1 (15.9, 20.5) |
| **Age, mean±SD** | 29.8±9.8 | 29.6, 30.1 | 29.8±9.7 | 29.6, 30.1 | 29.8±10.1 | 29.5, 30.2 |
| <20 years | 2,307 | 18.7 (17.9, 19.6) | 1,322 | 17.9 (16.9, 18.8) | 985 | 20 (18.7, 21.5) |
| 20-34 years | 5,770 | 46.8 (45.7, 48.0) | 3,580 | 48.3 (47.0, 49.6) | 2,190 | 44.6 (42.9, 46.3) |
| 35-49 years | 4,245 | 34.4 (33.4, 35.5) | 2,506 | 33.8 (32.7, 35.0) | 1,739 | 35.4 (33.7, 37.1) |
| **Marital status** |  |  |  |  |  |  |
| Unmarried | 3,400 | 27.6 (26.3, 28.9) | 1,632 | 22 (20.8, 23.4) | 1,768 | 36 (34.0, 38.0) |
| Married or living together | 8,633 | 70.1 (68.7, 71.4) | 5,532 | 74.7 (73.3, 76.0) | 3,101 | 63.1 (61.1, 65.1) |
| Divorced or not living together | 288 | 2.3 (2.06, 2.66) | 245 | 3.3 (2.88, 3.79) | 44 | 0.9 (0.65, 1.21) |
| **Wealth** |  |  |  |  |  |  |
| Poorest | 2,095 | 17 (15.1, 19.0) | 1,344 | 18.1 (16.1, 20.3) | 751 | 15.3 (13.5, 17.3) |
| Poorer | 2,304 | 18.7 (16.9, 20.7) | 1,371 | 18.5 (16.7, 20.5) | 933 | 19 (17.0, 21.2) |
| Middle | 2,468 | 20 (18.4, 21.8) | 1,511 | 20.4 (18.6, 22.3) | 957 | 19.5 (17.7, 21.3) |
| Richer | 2,839 | 23 (21.1, 25.1) | 1,704 | 23 (21.1, 25.0) | 1,135 | 23.1 (21.0, 25.4) |
| Richest | 2,616 | 21.2 (18.7, 24.0) | 1,479 | 20 (17.5, 22.7) | 1,137 | 23.1 (20.4, 26.1) |
| **Education** |  |  |  |  |  |  |
| No education | 2,335 | 19 (17.6, 20.4) | 1,942 | 26.2 (24.5, 27.9) | 393 | 8 (6.85, 9.34) |
| Basic | 4,155 | 33.7 (32.3, 35.2) | 2,256 | 30.5 (29.0, 32.0) | 1,898 | 38.6 (36.5, 40.8) |
| Secondary | 5,175 | 42 (40.3, 43.7) | 2,931 | 39.6 (37.7, 41.4) | 2,244 | 45.7 (43.7, 47.7) |
| Higher | 657 | 5.3 (4.56, 6.22) | 280 | 3.8 (3.11, 4.58) | 377 | 7.7 (6.50, 9.05) |
| **Occupation** |  |  |  |  |  |  |
| Not working | 2,704 | 21.9 (20.5, 23.4) | 2,032 | 27.4 (25.5, 29.4) | 672 | 13.7 (12.2, 15.3) |
| Agriculture | 4,747 | 38.5 (36.2, 40.9) | 3,591 | 48.5 (45.6, 51.3) | 1,156 | 23.5 (21.4, 25.8) |
| Professional/technical/ managerial/ clerical | 1,096 | 8.9 (8.06, 9.80) | 532 | 7.2 (6.35, 8.13) | 563 | 11.5 (10.1, 12.9) |
| Sales and service | 1,311 | 10.6 (9.70, 11.6) | 606 | 8.2 (7.23, 9.24) | 705 | 14.3 (12.9, 15.9) |
| Skilled/unskilled labor | 2,451 | 19.9 (18.6, 21.2) | 639 | 8.6 (7.58, 9.80) | 1,812 | 36.9 (34.7, 39.1) |
| Other | 13 | 0.1 (0.05, 0.22) | 8 | 0.1 (0.04, 0.28) | 6 | 0.1 (0.05, 0.29) |
| **Current smoke** |  |  |  |  |  |  |
|  | n | % (95%CI) | n | % (95%CI) | n | % (95%CI) |
| Do not smoke | 10,289 | 83.5 (82.5, 84.4) | 7,091 | 95.7 (95.0, 96.3) | 3,198 | 65.1 (63.0, 67.1) |
| Every day | 1,623 | 13.2 (12.4, 14.0) | 220 | 3 (2.49, 3.53) | 1,403 | 28.6 (26.7, 30.4) |
| Some days | 410 | 3.3 (2.93, 3.78) | 98 | 1.3 (1.00, 1.75) | 312 | 6.3 (5.51, 7.30) |
| **Current alcohol** |  |  |  |  |  |  |
| Never drinker | 1,716 | 13.9 (13.0, 15.0) | 940 | 12.7 (11.5, 13.9) | 776 | 15.8 (14.1, 17.6) |
| No drink in past month | 7,730 | 62.7 (61.0, 64.4) | 5,670 | 76.5 (74.7, 78.2) | 2,061 | 41.9 (39.2, 44.8) |
| Some drink in past month | 2,571 | 20.9 (19.7, 22.1) | 746 | 10.1 (9.01, 11.2) | 1,825 | 37.2 (35.0, 39.3) |
| Everyday drink | 304 | 2.5 (2.02, 3.01) | 53 | 0.7 (0.50, 1.01) | 252 | 5.1 (4.17, 6.27) |
| **Disability** |  |  |  |  |  |  |
| No difficulty | 9,921 | 80.5 (79.5, 81.5) | 5,826 | 78.7 (77.4, 79.9) | 4,094 | 83.4 (82.1, 84.6) |
| Some difficulty | 2,200 | 17.9 (17.0, 18.8) | 1,455 | 19.6 (18.5, 20.9) | 745 | 15.2 (14.0, 16.4) |
| A lot difficulty | 18 | 0.1 (0.09, 0.24) | 11 | 0.1 (0.07, 0.27) | 8 | 0.2 (0.08, 0.31) |
| Cannot do at all | 179 | 1.5 (1.24, 1.71) | 115 | 1.6 (1.29, 1.88) | 64 | 1.3 (1.00, 1.69) |
| Missing | 3 |  | 1 |  | 2 |  |
| **Health insurance** | 1,556 | 12.6 (11.3, 14.1) | 904 | 12.2 (10.8, 13.7) | 652 | 13.3 (11.7, 15.0) |

*n: frequency; %: percent; CI: confidence interval; SD: standard deviation*

The prevalence of mild, moderate, severely moderate and severe depression were 13.2% (95%CI: 12.3, 15.), 2.9% (95%CI: 2.5, 3.2), 0.9% (95%CI: 0.7, 1.1) and 0.2% (95%CI: 0.2, 0.4) respectively with the 4.0% (95%CI: 3.5, 4.5) overall prevalence of depression. The prevalence of mild to moderate anxiety and severe anxiety were 16.7% (95%CI: 15.6, 17.9) and 1.0% (95%CI: 0.8,1.2) respectively with the 17.7% (95%CI: 16.5, 18.9) overall prevalence of anxiety. Of total participants, 18.0% (95%CI: 16.8, 19.2) had depression or anxiety.

**Figure 1:**
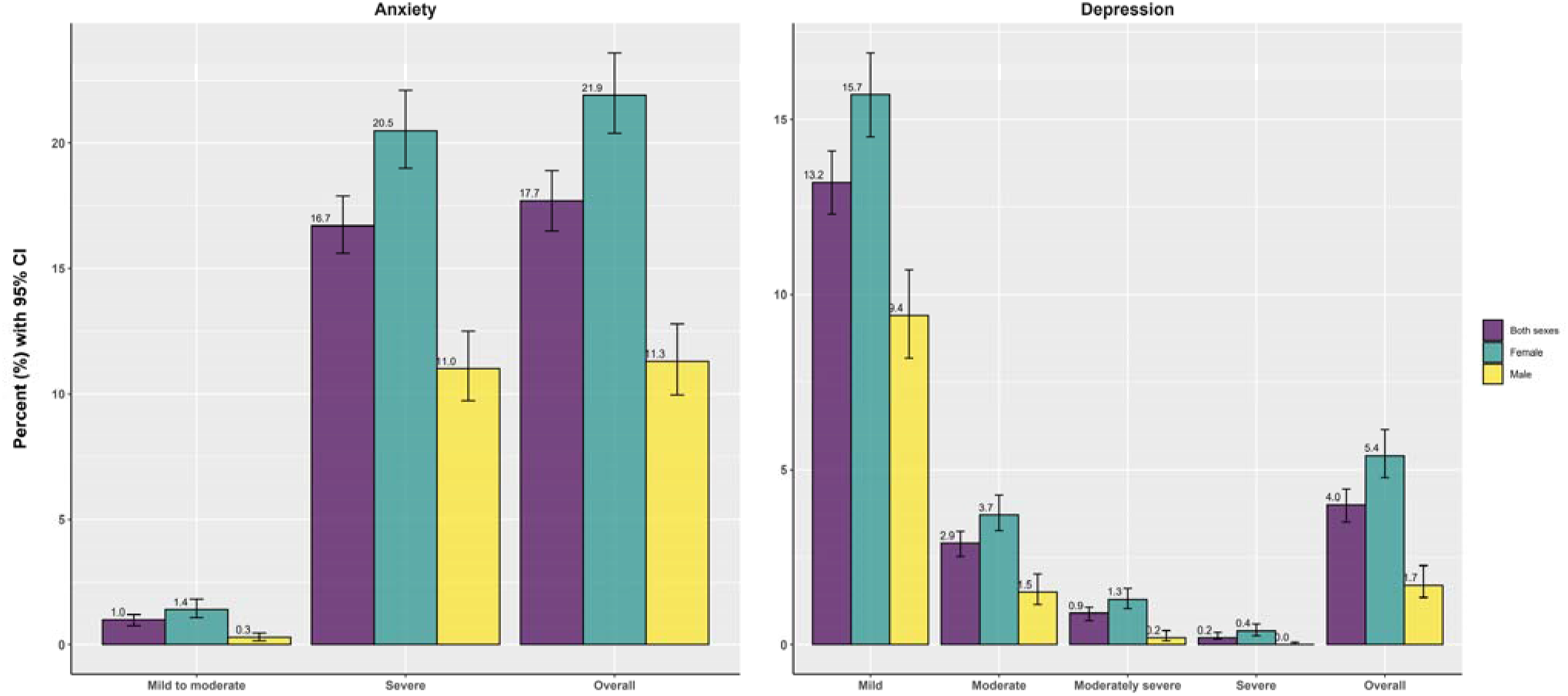
Gender wise prevalence of depression and anxiety among Nepalese population aged 15-49 years.

Sex, marital status, province, ethnicity, marital status, wealth quintile, alcohol use, and disability status were found to be associated with anxiety among participants. With female as reference, males had lower odds of developing anxiety (AOR=0.42, 95% CI: 0.36, 0.50). Participants who were divorced or not living together were found to have 2 folds higher odds (AOR=2.39, 95% CI: 1.73, 3.30) of developing anxiety. Among provinces, considering Koshi as reference, residents of Madhesh province had lower odds of having anxiety (AOR=0.63; 95% of CI: 0.45, 0.87), while no significant association was found with other provinces. Although residents of Gandaki Province were found to have lower odds of having anxiety in bivariate analysis (COR=0.67, 95% CI: 0.49, 0.93), multivariable analysis did not reveal any statistically significant association. Among different ethnic groups, Janajati had lower odds (AOR=0.77. 95% CI: 0.64, 0.91) of developing anxiety compared to Brahmin/Chhetri. Participants belonging to Dalit ethnic group were found to have higher odds (AOR=1.29, 95% CI:1.05, 1.59) of developing anxiety in multivariable regression model. Compared to participants in the poorest wealth quintile, participants in poorer wealth quintile had higher odds (AOR=1.20, 95% CI: 1.01, 1.43) of developing anxiety. Compared to those who never drink, participants who had ever drank but not in last one month were found to have lower odds (AOR=0.70, 95% CI: 0.59, 0.84) of developing anxiety. Regarding disability, compared to participants who did not have any difficulty, participants who had some difficulty (AOR: 1.82, 95% CI: 1.57, 2.10) and had lot of difficulty/can’t do at all (AOR: 2.10, 95% CI: 1.48, 2.97).

**Table 2:** Prevalence and factors associated with anxiety.

| Characteristics | % (95% CI) | COR (95%CI) | p-value | AOR (95%CI) | p-value |
| --- | --- | --- | --- | --- | --- |
| Age |  |  |  |  |  |
| <20 years | 14.8 (13.0, 16.7) | Ref |  | Ref |  |
| 20-34 years | 18.3 (16.8, 19.9) | **1.29 (1.11, 1.51)** | **0.001** | 1.12 (0.90, 1.41) | 0.313 |
| 35-49 years | 18.4 (16.9, 20.1) | **1.31 (1.11, 1.53)** | **0.001** | 0.94 (0.72, 1.23) | 0.653 |
| **Sex** |  |  |  |  |  |
| Female | 21.9 (20.4, 23.6) | **Ref** |  | Ref |  |
| Male | 11.3 (1.0, 12.8) | **0.45 (0.39, 0.53)** | **<0.001** | **0.42 (0.36, 0.50)** | **<0.001** |
| **Marital status** |  |  |  |  |  |
| Unmarried | 13.9 (12.5, 15.5) | **Ref** |  | Ref |  |
| Married /living together | 18.5 (17.2, 19.8) | **1.4 (1.23, 1.60)** | **<0.001** | 1.13 (0.94, 1.36) | 0.206 |
| Divorced/ separated | 39 (32.9, 45.5) | **3.96 (2.99, 5.25)** | **<0.001** | **2.39 (1.73, 3.30)** | **<0.001** |
| **Place of residence** |  |  |  |  |  |
| Urban | 17.6 (16.1, 19.2) | Ref |  | Ref |  |
| Rural | 17.9 (16.4, 19.6) | 1.03 (0.88, 1.19) | 0.743 | 0.97 (0.82, 1.13) | 0.675 |
| **Ecological belt** |  |  |  |  |  |
| Mountain | 20.3 (16.3, 24.9) | Ref |  | Ref |  |
| Hill | 16.2 (14.8, 17.7) | 0.76 (0.57, 1.01) | 0.06 | 0.91 (0.71, 1.18) | 0.473 |
| Terai | 18.5 (16.8, 20.4) | 0.89 (0.67, 1.19) | 0.437 | 1.24 (0.92, 1.69) | 0.159 |
| **Province** |  |  |  |  |  |
| Koshi | 20 (17.1, 23.1) | Ref |  | Ref |  |
| Madhesh | 15.9 (13.0, 19.2) | 0.76 (0.56, 1.02) | 0.065 | **0.63 (0.45, 0.87)** | **0.005** |
| Bagmati | 16.5 (14.4, 18.9) | 0.79 (0.62, 1.02) | 0.069 | 1 (0.78, 1.30) | 0.975 |
| Gandaki | 14.4 (11.4, 18.0) | **0.67 (0.49, 0.93)** | **0.017** | 0.76 (0.55, 1.05) | 0.092 |
| Lumbini | 18.1 (15.3, 21.3) | 0.88 (0.67, 1.17) | 0.383 | 0.77 (0.57, 1.02) | 0.070 |
| Karnali | 24.1 (20.9, 27.7) | 1.28 (0.98, 1.67) | 0.071 | 1.28 (0.97, 1.70) | 0.080 |
| Sudurpashchim | 18.6 (15.9, 21.7) | 0.92 (0.70, 1.20) | 0.531 | 0.81 (0.61, 1.08) | 0.150 |
| **Ethnicity** |  |  |  |  |  |
| Brahmin/Chhetri | 18.1 (16.4, 20.0) | Ref |  | Ref |  |
| Dalit | 23.5 (20.5, 26.8) | **1.39 (1.14, 1.70)** | **0.001** | **1.29 (1.05, 1.59)** | **0.014** |
| Janajati | 15.9 (14.6, 17.3) | **0.86 (0.74, 0.99)** | **0.041** | **0.77 (0.64, 0.91)** | **0.003** |
| Madheshi | 16.6 (14.0, 19.6) | 0.9 (0.71, 1.14) | 0.381 | 1.06 (0.81, 1.40) | 0.666 |
| Others | 15.3 (11.2, 20.5) | 0.81 (0.56, 1.19) | 0.291 | 0.99 (0.64, 1.53) | 0.961 |
| **Religion** |  |  |  |  |  |
| Hindu | 18 (16.7, 19.3) | Ref |  | Ref |  |
| Other | 16.3 (14.3, 18.4) | 0.89 (0.75, 1.04) | 0.139 | 0.99 (0.80, 1.23) | 0.925 |
| **Wealth** |  |  |  |  |  |
| Poorest | 19 (17.1, 21.0) | Ref |  | Ref |  |
| Poorer | 20.6 (18.3, 23.0) | 1.11 (0.93, 1.32) | 0.252 | **1.20 (1.01, 1.43)** | **0.034** |
| Middle | 18.5 (16.5, 20.6) | 0.97 (0.81, 1.15) | 0.702 | 1.02 (0.84, 1.25) | 0.816 |
| Richer | 17.2 (15.3, 19.2) | 0.89 (0.74, 1.06) | 0.177 | 0.97 (0.78, 1.21) | 0.770 |
| Richest | 13.9 (11.8, 16.3) | **0.69 (0.55, 0.86)** | **0.001** | 0.8 (0.60, 1.08) | 0.140 |
| **Education** |  |  |  |  |  |
| no education | 22.1 (19.7, 24.8) | Ref |  | Ref |  |
| Basic | 18.2 (16.6, 19.9) | **0.78 (0.67, 0.92)** | **0.003** | 1.02 (0.86, 1.21) | 0.836 |
| Secondary | 15.8 (14.5, 17.2) | **0.66 (0.56, 0.78)** | **<0.001** | 1 (0.83, 1.22) | 0.961 |
| Higher | 13.2 (10.1, 17.2) | **0.54 (0.38, 0.76)** | **<0.001** | 0.99 (0.68, 1.43) | 0.938 |
| **Occupation** |  |  |  |  |  |
| Not working | 17.5 (15.5, 19.6) | Ref |  | Ref |  |
| Agriculture | 20.3 (18.7, 22.0) | 1.21 (1.04, 1.40) | 0.014 | 1 (0.85, 1.19) | 0.962 |
| Professional/technical/<br>managerial/ clerical | 15.4 (12.8, 18.3) | 0.86 (0.67, 1.09) | 0.214 | 1.01 (0.78, 1.31) | 0.930 |
| Sales and service | 14 (11.9, 16.5) | 0.77 (0.61, 0.97) | 0.026 | 0.87 (0.67, 1.11) | 0.261 |
| Skilled/unskilled labor | 15.8 (13.7, 18.1) | 0.89 (0.73, 1.08) | 0.225 | 1.01 (0.82, 1.25) | 0.898 |
| Other | 21.1 (5.07, 57.3) | 1.27 (0.25, 6.36) | 0.774 | 1.38 (0.25, 7.49) | 0.708 |
| **Current smoke** |  |  |  |  |  |
| Do not smoke | 18.1 (16.8, 19.5) | Ref |  | Ref |  |
| every day | 14.3 (12.3, 16.5) | **0.75 (0.63, 0.91)** | **0.003** | 0.95 (0.76, 1.18) | 0.617 |
| some days | 20.1 (15.7, 25.4) | 1.14 (0.84, 1.55) | 0.398 | 1.36 (0.97, 1.91) | 0.074 |
| **Current alcohol** |  |  |  |  |  |
| Never drinker | 19.4 (17.3, 21.7) | Ref |  | Ref |  |
| No drink in past month | 17.8 (16.4, 19.4) | 0.9 (0.76, 1.06) | 0.196 | **0.7 (0.59, 0.84)** | **<0.001** |
| Some drink in past month | 15.9 (14.0, 17.9) | **0.78 (0.64, 0.95)** | **0.016** | 0.94 (0.77, 1.16) | 0.592 |
| Everyday drink | 19.7 (14.5, 26.1) | 1.02 (0.69, 1.50) | 0.934 | 1.2 (0.82, 1.77) | 0.35 |
| **Disability** |  |  |  |  |  |
| No difficulty | 15.4 (14.3, 16.6) | Ref |  | Ref |  |
| Some difficulty | 26.8 (24.3, 29.4) | **2.01 (1.75, 2.29)** | **<0.001** | **1.82 (1.57, 2.10)** | **<0.001** |
| A lot difficulty to cannot do at all | 29.1 (22.8, 36.4) | **2.25 (1.61, 3.14)** | **<0.001** | **2.10 (1.48, 2.97)** | **<0.001** |

n: frequency; %: percent; CI: confidence interval; COR: crude odds ratio; AOR: adjusted odds ratio; Ref: reference group **Bold** represents significance at 5% level of significance.

The presence of depression among participants was found to be associated with various factors including sex, marital status, province, ethnicity, alcohol use, and disability status. Individuals who were divorced or separated had three times higher odds (AOR=3.11, 95% CI: 1.81, 5.35) of developing depression. Compared to females, males were found to have three folds lower odds (AOR=**0.29, 95% CI: 0.21, 0.39)** of having depression, In the bivariate analysis, residents of although residents of Gandaki province were found to have lower odds of having depression in bivariate analysis (COR=0.65, 95% CI: 0.42, 0.99), and residents in Karnali province were found to have higher odds (COR=1.56, 95% CI: 1.13, 2.16) of developing depression, multivariable analysis did not reveal any statistically significant association. Among different ethnic groups, Janajati had lower odds (AOR=0.67, 95% CI: 0.48, 0.92) of developing depression compared to Brahmin/Chhetri. Participants who had previously consumed alcohol but not in the past month had lower odds (AOR=0.59, 95% CI: 0.43, 0.80) of developing anxiety compared to those who had never consumed alcohol. Additionally, participants who faced some difficulty had higher odds (AOR: 1.96, 95% CI: 1.52, 2.51) of developing depression compared to those without any difficulty.

**Table 3:**
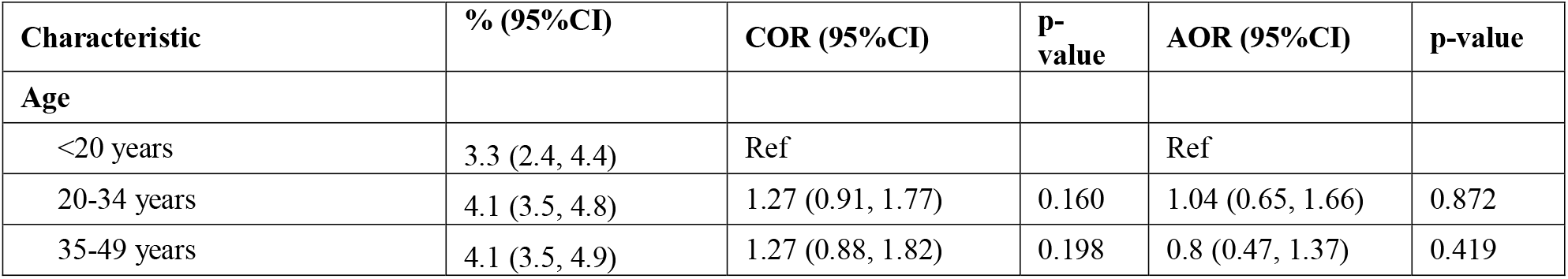
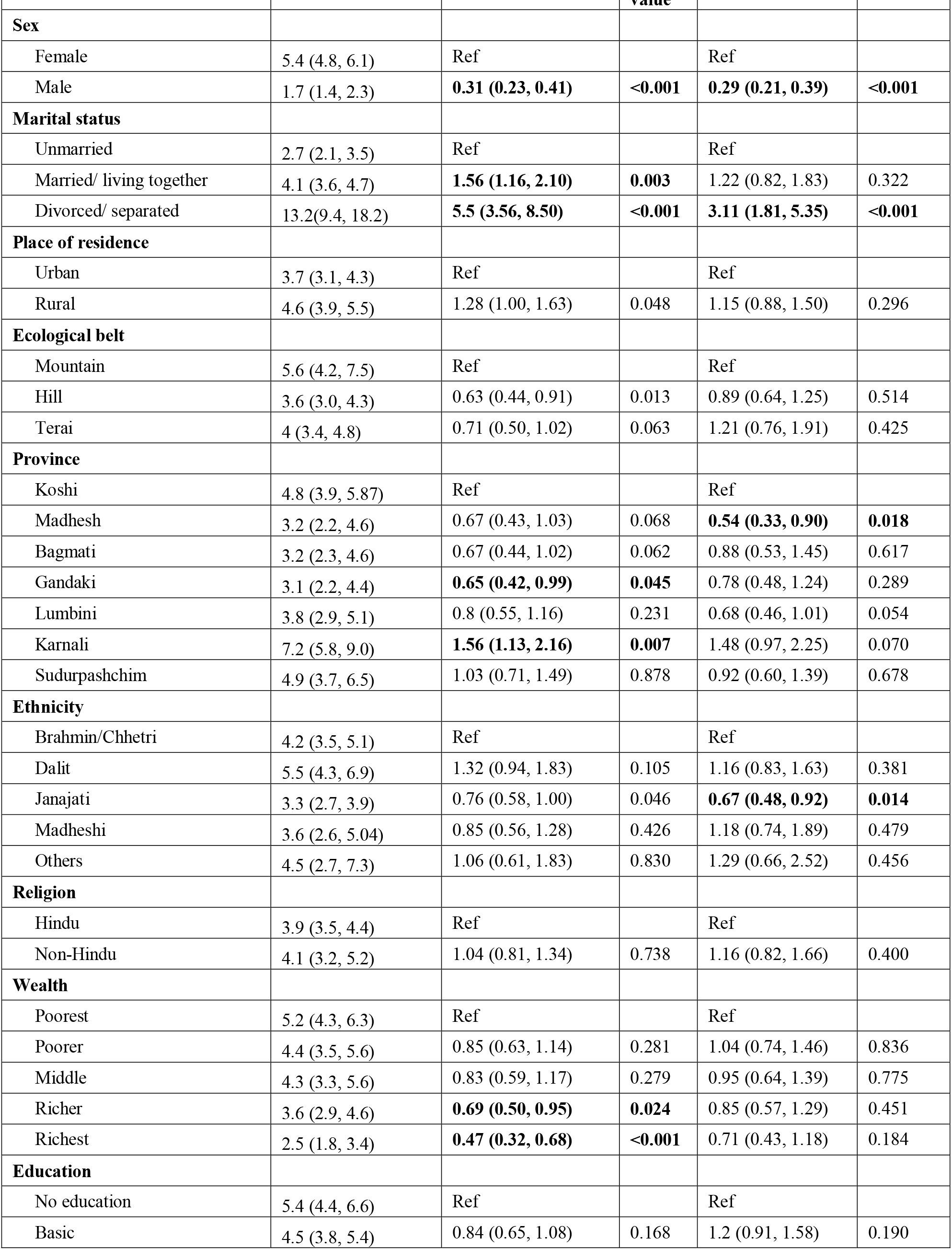
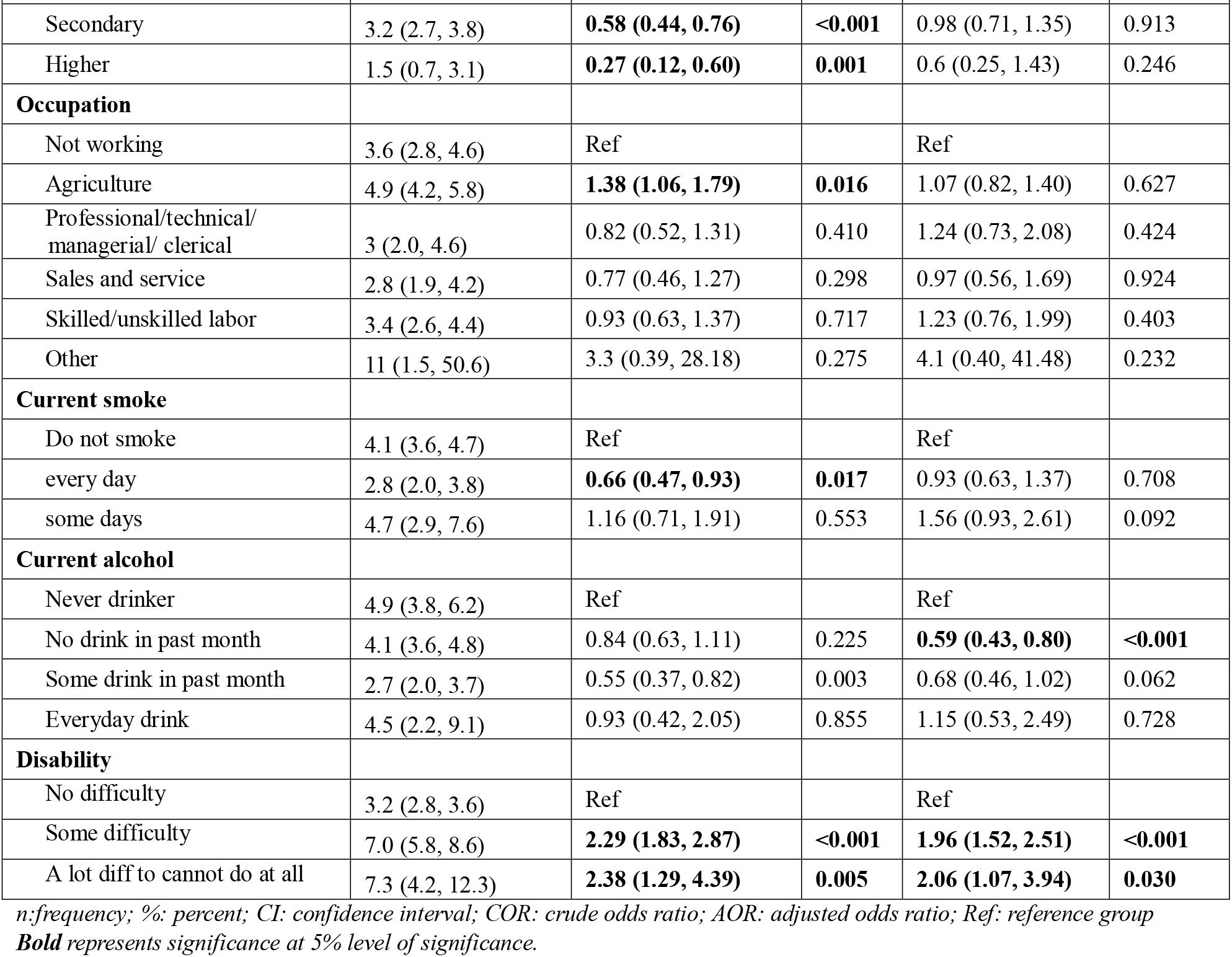
Prevalence and factors associated with depression.

| Characteristic | % (95%CI) | COR (95%CI) | P-value | AOR (95%CI) | p-value |
| --- | --- | --- | --- | --- | --- |
| **Age** |  |  |  |  |  |
| <20 years | 3.3 (2.4, 4.4) | Ref |  | Ref |  |
| 20-34 years | 4.1 (3.5, 4.8) | 1.27 (0.91, 1.77) | 0.160 | 1.04 (0.65, 1.66) | 0.872 |
| 35-49 years | 4.1 (3.5, 4.9) | 1.27 (0.88, 1.82) | 0.198 | 0.8 (0.47, 1.37) | 0.419 |
| **Sex** |  |  |  |  |  |
| Female | 5.4 (4.8, 6.1) | Ref |  | Ref |  |
| Male | 1.7 (1.4, 2.3) | **0.31 (0.23, 0.41)** | **<0.001** | **0.29 (0.21, 0.39)** | **<0.001** |
| **Marital status** |  |  |  |  |  |
| Unmarried | 2.7 (2.1, 3.5) | Ref |  | Ref |  |
| Married/ living together | 4.1 (3.6, 4.7) | **1.56 (1.16, 2.10)** | **0.003** | 1.22 (0.82, 1.83) | 0.322 |
| Divorced/ separated | 13.2(9.4, 18.2) | **5.5 (3.56, 8.50)** | **<0.001** | **3.11 (1.81, 5.35)** | **<0.001** |
| **Place of residence** |  |  |  |  |  |
| Urban | 3.7 (3.1, 4.3) | Ref |  | Ref |  |
| Rural | 4.6 (3.9, 5.5) | 1.28 (1.00, 1.63) | 0.048 | 1.15 (0.88, 1.50) | 0.296 |
| **Ecological belt** |  |  |  |  |  |
| Mountain | 5.6 (4.2, 7.5) | Ref |  | Ref |  |
| Hill | 3.6 (3.0, 4.3) | 0.63 (0.44, 0.91) | 0.013 | 0.89 (0.64, 1.25) | 0.514 |
| Terai | 4 (3.4, 4.8) | 0.71 (0.50, 1.02) | 0.063 | 1.21 (0.76, 1.91) | 0.425 |
| **Province** |  |  |  |  |  |
| Koshi | 4.8 (3.9, 5.87) | Ref |  | Ref |  |
| Madhesh | 3.2 (2.2, 4.6) | 0.67 (0.43, 1.03) | 0.068 | **0.54 (0.33, 0.90)** | **0.018** |
| Bagmati | 3.2 (2.3, 4.6) | 0.67 (0.44, 1.02) | 0.062 | 0.88 (0.53, 1.45) | 0.617 |
| Gandaki | 3.1 (2.2, 4.4) | **0.65 (0.42, 0.99)** | **0.045** | 0.78 (0.48, 1.24) | 0.289 |
| Lumbini | 3.8 (2.9, 5.1) | 0.8 (0.55, 1.16) | 0.231 | 0.68 (0.46, 1.01) | 0.054 |
| Karnali | 7.2 (5.8, 9.0) | **1.56 (1.13, 2.16)** | **0.007** | 1.48 (0.97, 2.25) | 0.070 |
| Sudurpashchim | 4.9 (3.7, 6.5) | 1.03 (0.71, 1.49) | 0.878 | 0.92 (0.60, 1.39) | 0.678 |
| **Ethnicity** |  |  |  |  |  |
| Brahmin/Chhetri | 4.2 (3.5, 5.1) | Ref |  | Ref |  |
| Dalit | 5.5 (4.3, 6.9) | 1.32 (0.94, 1.83) | 0.105 | 1.16 (0.83, 1.63) | 0.381 |
| Janajati | 3.3 (2.7, 3.9) | 0.76 (0.58, 1.00) | 0.046 | **0.67 (0.48, 0.92)** | **0.014** |
| Madheshi | 3.6 (2.6, 5.04) | 0.85 (0.56, 1.28) | 0.426 | 1.18 (0.74, 1.89) | 0.479 |
| Others | 4.5 (2.7, 7.3) | 1.06 (0.61, 1.83) | 0.830 | 1.29 (0.66, 2.52) | 0.456 |
| **Religion** |  |  |  |  |  |
| Hindu | 3.9 (3.5, 4.4) | Ref |  | Ref |  |
| Non-Hindu | 4.1 (3.2, 5.2) | 1.04 (0.81, 1.34) | 0.738 | 1.16 (0.82, 1.66) | 0.400 |
| **Wealth** |  |  |  |  |  |
| Poorest | 5.2 (4.3, 6.3) | Ref |  | Ref |  |
| Poorer | 4.4 (3.5, 5.6) | 0.85 (0.63, 1.14) | 0.281 | 1.04 (0.74, 1.46) | 0.836 |
| Middle | 4.3 (3.3, 5.6) | 0.83 (0.59, 1.17) | 0.279 | 0.95 (0.64, 1.39) | 0.775 |
| Richer | 3.6 (2.9, 4.6) | **0.69 (0.50, 0.95)** | **0.024** | 0.85 (0.57, 1.29) | 0.451 |
| Richest | 2.5 (1.8, 3.4) | **0.47 (0.32, 0.68)** | **<0.001** | 0.71 (0.43, 1.18) | 0.184 |
| **Education** |  |  |  |  |  |
| No education | 5.4 (4.4, 6.6) | Ref |  | Ref |  |
| Basic | 4.5 (3.8, 5.4) | 0.84 (0.65, 1.08) | 0.168 | 1.2 (0.91, 1.58) | 0.190 |
| Secondary | 3.2 (2.7, 3.8) | **0.58 (0.44, 0.76)** | **<0.001** | 0.98 (0.71, 1.35) | 0.913 |
| Higher | 1.5 (0.7, 3.1) | **0.27 (0.12, 0.60)** | **0.001** | 0.6 (0.25, 1.43) | 0.246 |
| **Occupation** |  |  |  |  |  |
| Not working | 3.6 (2.8, 4.6) | Ref |  | Ref |  |
| Agriculture | 4.9 (4.2, 5.8) | **1.38 (1.06, 1.79)** | **0.016** | 1.07 (0.82, 1.40) | 0.627 |
| Professional/technical/<br>managerial/ clerical | 3 (2.0, 4.6) | 0.82 (0.52, 1.31) | 0.410 | 1.24 (0.73, 2.08) | 0.424 |
| Sales and service | 2.8 (1.9, 4.2) | 0.77 (0.46, 1.27) | 0.298 | 0.97 (0.56, 1.69) | 0.924 |
| Skilled/unskilled labor | 3.4 (2.6, 4.4) | 0.93 (0.63, 1.37) | 0.717 | 1.23 (0.76, 1.99) | 0.403 |
| Other | 11 (1.5, 50.6) | 3.3 (0.39, 28.18) | 0.275 | 4.1 (0.40, 41.48) | 0.232 |
| **Current smoke** |  |  |  |  |  |
| Do not smoke | 4.1 (3.6, 4.7) | Ref |  | Ref |  |
| every day | 2.8 (2.0, 3.8) | **0.66 (0.47, 0.93)** | **0.017** | 0.93 (0.63, 1.37) | 0.708 |
| some days | 4.7 (2.9, 7.6) | 1.16 (0.71, 1.91) | 0.553 | 1.56 (0.93, 2.61) | 0.092 |
| **Current alcohol** |  |  |  |  |  |
| Never drinker | 4.9 (3.8, 6.2) | Ref |  | Ref |  |
| No drink in past month | 4.1 (3.6, 4.8) | 0.84 (0.63, 1.11) | 0.225 | **0.59 (0.43, 0.80)** | **<0.001** |
| Some drink in past month | 2.7 (2.0, 3.7) | 0.55 (0.37, 0.82) | 0.003 | 0.68 (0.46, 1.02) | 0.062 |
| Everyday drink | 4.5 (2.2, 9.1) | 0.93 (0.42, 2.05) | 0.855 | 1.15 (0.53, 2.49) | 0.728 |
| **Disability** |  |  |  |  |  |
| No difficulty | 3.2 (2.8, 3.6) | Ref |  | Ref |  |
| Some difficulty | 7.0 (5.8, 8.6) | **2.29 (1.83, 2.87)** | **<0.001** | **1.96 (1.52, 2.51)** | **<0.001** |
| A lot diff to cannot do at all | 7.3 (4.2, 12.3) | **2.38 (1.29, 4.39)** | **0.005** | **2.06 (1.07, 3.94)** | **0.030** |

*n*:frequency; %: percent; CI: confidence interval; COR: crude odds ratio; AOR: adjusted odds ratio; Ref: reference group
**Bold** represents significance at 5% level of significance.

### Care seeking behavior for anxiety and depression

Of 2217 participants with depression or anxiety in the past 2 weeks, 32.9% (95% CI: 30.4, 34.4) tried to seek help for the things they experienced. Care seeking among males and females were 29.2% (24.8, 34.2) and 34.1% (31.3, 37.0). Among those who seek care, most of the participants seek care from family member other than spouse (44.6%, 95%CI: 40.5, 48.6), from friends(37.0%; 95%CI: 33.0, 41.2), spouse(26.6%, 95%CI: 22.9, 30.7), neighbor (10.9%; 95% CI: 8.8, 13.5), health care providers (9.4%; 95%CI: 7.3, 12.0) and least seek care from social workers and community health workers (<1%).

**Table 4:** Care seeking behavior among participants with anxiety and depression.

| Care seeking from # | Overall, n= 728 |  | Female, n = 563 |  | Male, n = 166 |  |
| --- | --- | --- | --- | --- | --- | --- |
|  | n (%) | 95% CI | n (%) | 95% CI | n (%) | 95% CI |
| Among participants having either |  |  |  |  |  |  |
| **anxiety or depression** |  |  |  |  |  |  |
| Family members except spouse | 325 (44.6) | 40.5, 48.6 | 256 (45.6) | 41.0, 50.2 | 68 (41.1) | 33.1, 49.6 |
| Friend | 270 (37.0) | 33.0, 41.2 | 184 (32.7) | 28.1, 37.6 | 86 (51.7) | 43.8, 59.6 |
| Spouse | 194 (26.6) | 22.9, 30.7 | 165 (29.3) | 25.0, 34.1 | 29 (17.5) | 11.6, 25.4 |
| Neighbour | 79 (10.9) | 8.77, 13.5 | 64 (11.3) | 8.82, 14.4 | 16 (9.6) | 6.16, 14.6 |
| Health care provider | 68 (9.4) | 7.30, 12.0 | 52 (9.2) | 6.81, 12.4 | 17 (10.0) | 6.23, 15.7 |
| Social worker | 2 (0.3) | 0.07, 1.37 | 0 (0.0) |  | 2.0 (1.3) | 0.30, 5.89 |
| Community health workers | 2 (0.2) | 0.08, 0.79 | 2 (0.3) | 0.10, 1.02 | 0 (0.0) |  |
| Religious leader | 3 (0.4) | 0.13, 1.39 | 3 (0.5) | 0.17, 1.79 | 0 (0.0) |  |
| **Among participants having depression** |  |  |  |  |  |  |
| Family members except spouse | 104 (50.0) | 43.0, 57.0 | 86 (48.7) | 41.0, 56.5 | 18 (57.1) | 39.2, 73.4 |
| Spouse | 53 (25.4) | 18.9, 33.1 | 48 (27.3) | 19.8, 36.3 | 5 (15.0) | 6.51, 31.0 |
| Friend | 57 (27.5) | 21.3, 34.8 | 46 (26.3) | 19.6, 34.3 | 11 (34.2) | 19.6, 52.6 |
| Health care provider | 30 (14.5) | 9.68, 21.2 | 27 (15.1) | 9.65, 22.9 | 4 (11.2) | 4.14, 26.8 |
| Neighbour | 25 (11.9) | 8.18, 17.0 | 21 (11.9) | 7.69, 17.8 | 4 (12.3) | 5.73, 24.3 |
| Social worker |  |  |  |  |  |  |
| Religious leader | 2 (1.2) | 0.27, 4.71 | 2 (1.4) | 0.33, 5.53 | 0 (0.0) |  |
| Community health workers | 0 (0.2) | 0.03, 1.57 | 0 (0.3) | 0.04, 1.86 | 0 (0.0) |  |
| **Among participants having anxiety** |  |  |  |  |  |  |
| Family members except spouse | 319 (44.5) | 40.4, 48.6 | 252 (45.5) | 40.9, 50.1 | 67 (41.2) | 33.1, 49.8 |
| Friend | 263 (36.7) | 32.8, 40.8 | 180 (32.5) | 28.0, 37.3 | 83 (51.0) | 43.0, 58.9 |
| Spouse | 191 (26.7) | 22.9, 30.8 | 162 (29.3) | 25.0, 34.1 | 29 (17.7) | 11.8, 25.8 |
| Neighbor | 78 (10.9) | 8.70, 13.5 | 62 (11.2) | 8.70, 14.3 | 16 (9.7) | 6.25, 14.8 |
| Health care provider | 67 (9.4) | 7.25, 12.0 | 50 (9.1) | 6.70, 12.3 | 17 (10.2) | 6.34, 15.9 |
| Social worker | 2 (0.3) | 0.07, 1.39 | 0 (0.0) |  | 2 (1.4) | 0.30, 5.98 |
| Community health workers | 2 (0.3) | 0.08, 0.80 | 2 (0.3) | 0.10, 1.04 | 0 (0.0) |  |
| Religious leader | 2 (0.3) | 0.07, 1.26 | 2 (0.4) | 0.09, 1.63 | 0 (0.0) |  |

## Discussion

In our study, 11.3% of participants were found to have anxiety while 1.7% were found to have depression. The prevalence of depression in our study is notably lower than the pooled estimate of depression in 30 countries estimated from 68 studies.^20^ using the random-effect model was 12.9%. ^20^ using the random-effect model was 12.9%. The prevalence of depression varies by continent, with South America having the highest overall rate of 20.6%. The prevalence was found to be 16.7% Asia, 13.4% in North America, 11.9% in Europe and 11.5% in Africa. In comparison, Australia has the lowest rate of depression at 7.3%.^20^

### Sex differences

Our study revealed that females have higher odds of developing anxiety and depression compared to males. In one of previous systematic review that computed the pooled prevalence of depression in 30 countries from 68 studies, the prevalence of depression was found to be 14.4% among female and 11.5% among male.^20^ One other study has also indicated that women bear higher burden of anxiety^4^ and depression disorder.^21, 22^ A multi-country study has demonstrated that females were found to have 1.6 times in prevalence and DALYs of anxiety disorder compared to males globally.^4^

Although the reason for higher prevalence of anxiety disorder among women is not clearly understood, some factors more sensitivity to criticism, rejection, and separation ^20, 23, 24^, more frequent encounter of adverse life events such as sexual violence and harassment and higher rates of re-victimization could be responsible for higher prevalence of anxiety among females.^4, 23, 25–27^

Genetic factors, postnatal stress, cultural environment with unequal gender roles could have some role in exacerbating anxiety among females.^4, 5^ Depression is more common during pregnancy and among women who had recently given birth affecting over 10% of women in this group, which could be one factor for higher prevalence among women.^1, 28^ Some of the previous studies have suggested the onset of puberty may trigger a genetic susceptibility in females.^23^ Adolescent girls encounter more stress, particularly interpersonal stress, is known to play a role in the higher rates of depression seen could also be responsible for higher prevalence of depression among females.^23, 29^ The study findings indicate that women may need more specific and targeted interventions so as to reduce the burden of anxiety and depression at national and subnational level.

### Provincial differences

Residents in Madhesh province had relatively lower odds of having anxiety and depression compared to Koshi while Karnali province reported slightly higher rates. In one of the previous study, although anxiety and depression were not specifically assessed, Madhesh province was found to have substantially lower prevalence of lifetime mental disorder in one of the nationwide study in 2019-2020.^30^ Relatively lower prevalence of anxiety, depression or other mental disorder in Madhesh Province and higher prevalence in residents of Karnali province is not clearly understood. However, these differences could be because of some cultural practices, social immersion, cohesion and gathering which have to be further explored through qualitative studies.^30^

### Wealth quintile

Although no significant difference was noted based on wealth quintile on prevalence of depression, the participants belonging to poorer wealth quintile were found to have higher odds of having anxiety compared to the participants in poorest wealth quintile. In the bivariate analysis, participants in richest wealth quintile were found to have lower odds of developing anxiety as well as depression which did not prevail in the multivariable analysis. Although not every study had a statistically significant association between wealth and depression, the majority of studies included in the review exhibited a consistent pattern demonstrating inverse relation between wealth and depression.^31^ Multiple studies suggest that wealth provides a stronger protection against life adversities, provides a sense of financial stability, help manage family expectation and relationships which reduces the chances of life stressors^32–34^ and serves as buffer against anxiety and depression symptoms. ^31–34^

### Age

Although no significant difference was found in prevalence of depression, participants of age 20-34 years and 35-49 years were found to have slightly higher odds of having anxiety compared to participants below the age of 20 years. This could be because of some of the stressful life events like family separation, that people often encounter after age of 20 years in Nepalese context.

### Disability

Our study indicates that people with disabilities have higher odds of having anxiety and depression. Depression is an independent risk factor for disability (in old age) and disability increases the risk of depression indicating bi-directional relationship.^35^ Multiple other studies have shown that disability is associated with anxiety^36, 37^ and depression.^38, 39^ Although the direct link between depression is not clearly understood, depression is linked to particular life situations that are more common among people with disabilities. Furthermore, persons with disabilities face a variety of unique concerns and challenges that may chances of developing depression.^40^ Disability may mean having difficulties in walking around, doing daily works independently like bathing, that may often make them feel frustrated and embarrassed.^40^ People with disabilities often face the social barriers and isolation because of difficulties in joining social functions and gathering, forming social relationship and may often receive limited social support that puts them at higher risk of depression.^40^ Physical or other forms of limitation often puts them on higher risk of being unemployed because of social prejudice and misconception regarding disability.^40^

### Care seeking behaviour

Similar to other mental disorders, there is high undertreatment rate for anxiety and depression. In one of the previous study in Sweden, 40.9% of participants with depression, 36.8% of participants with anxiety and 60.9% of participants facing comorbid condition of anxiety and depression were found to seek care. ^41^ A systematic review involved analysis of 149 studies from 84 countries between 2000 to 2021, estimated that 33% of patients with major depressive disorder seek care in high income countries while the proportion was 15% in upper middle-income countries and only 8% in LMICs which highlight the notable proportion of the unmet need for mental health services.^42^ Apart from notably lower proportion of patients who seek care, delay in seeking care is other important factor that undermine the health outcomes among individuals with anxiety and depression. In the study in eleven countries: United States, United Kingdom, Italy, South Africa, Switzerland, Austria, Sweden, France, Argentina, Belgium and Venezuela, 40% of participants were found to seek care during the same year of encountering disorder while for other remaining participants, the median delay in seeking care from onset of symptoms to seeking care was 8 years.

The unwillingness or inability to seek assistance can be attributed to a range of factors, including high expenses, poor service quality, and limited resource availability. It might also be due to a lack of awareness on mental health, the existence of societal stigma associated with seeking therapy, and negative past experiences with seeking assistance.^1^ Further, individuals with mental problems are prevented from receiving the essential treatment because of a number of reasons at the individual, provider, and systemic levels. At the individual level, barriers like reluctance to disclose their issues, anxiety about stigma, time restraints, unfavorable views of treatment, cultural influences, a propensity to manage mental health issues alone, and a low willingness to embrace change all contribute to impeding their desire to receive treatment.^43^ The proper treatment of patients with anxiety and depression, is hampered at the provider level by a number of variables. These challenges include under-detection at the primary level, care professionals’ little familiarity with mental illnesses, and patients’ physical symptom presentation.^43^ A dearth of specialized mental health services, a paucity of health workers educated in anxiety and depression diagnosis and treatment techniques, and the absence of integration of mental health care into primary healthcare settings are system-level problems.^43^

### Health impacts, policy and programme implication

Anxiety disorders are consistently linked to significant impairments in both productive roles (such as work absenteeism, work performance, unemployment, and underemployment) and social roles (such as social isolation, interpersonal tensions, and marital disruption) in epidemiological surveys.^5^ Depression can have significant impact on the economy of the country. For example, the health and economic burden of depression was estimated to represent 2.9% of gross domestic products in Singapore.^44^

A substantial proportion of individuals with anxiety and depression do not seek medical attention in earlier stage. Further, health systems, particularly in low- and middle-income countries are less prepared for delivery of mental health service and are often underfunded.^5^ Depression is the leading risk factor for suicide with the problem being further exacerbated by substance use disorders.^5^ To transform the mental health agenda, it is critical to invest in addressing the fundamental social and economic factors that affect people’s mental well-being in addition to expanding access to high-quality services and care.^1^ According to WHO expanding the provision of treatment for depression and anxiety yields a benefit-to-cost ratio of 5 to 1 meaning that for every one dollar spent in treatment for depression and anxiety, there would be benefits equal to 5 dollar.^1^

Some of the strategies for expanding coverage of preventive and curative services included include school-based social and emotional learning programmes, integrating mental health services in primary health care connected with appropriate referral network with higher level facilities, a ban on the use of very toxic pesticides (to prevent suicides), and improved availability of treatment provisions specified in the WHO UHC Compendium.^1^ individuals with anxiety and depression can benefit from community-based mental health care because such services are accessible to people.^1^ Frequent co-occurance of anxiety and depression and their bidirectional association with conditions like obesity, and chronic conditions like type 2 diabetes mellitus, coronary artery disease, and chronic pain disorders ^5^ also point out the need of integrated care.

Task-sharing with primary healthcare practitioners has been shown to be successful in reducing the treatment gap and increasing coverage for priority mental health problems^1^ which could be particularly relevant for countries like Nepal facing dearth of psychiatrist and specialized health care facilities for treatment of mental disorder. More people with depression seek support from friends than spouse for anxiety and depression and thus peer support programmes, where people draw on their personal experiences to help one another could be useful in Nepalese context. WHO suggest that peer support programmes could help in information exchange, advocacy and increasing awareness, providing emotional support, providing helpful assistance, creating social contacts.^1^

### Strengths and limitations of the study

Most of previous studies on anxiety and depression in Nepal are confined to specific group of participants like health care workers during COVID-19 pandemic ^10, 11^, nurses^12, 13^, traffic police^14^, among patients of type II diabetes^15, 16^, people living in quarantine centre during COVID-19 pandemic^45^, patients of MDR TB^17^ and hypertensive adults^18^ and are confined in specific localities. However, our study involves an analysis from nationally representative sample taking into consideration the current federal structure. However, despite being one of the large-scale studies in nationally representative sample, our study has some limitations. The study was not primarily designed to study anxiety and depression but was a part of more comprehensive study that assessed multiple other health problems like reproductive, maternal, newborn and child health, fertility behavior, hypertension etc. So, some of the variables that could be important like stress coping skills, meditation behaviour, social capital and social support which could be potentially associated with anxiety and depression have not been assessed in this study. ^10, 11^.

## Conclusion

The prevalence of depression and anxiety is relatively higher among females compared to males. Depression was associated negatively with the Janajati ethnic group, males and Madhesh province whereas positively associated with divorced or separated or not living together status of marriage. Anxiety was associated positively with Dalit ethnic group, poorest wealth quintile, divorced or separated or not living together status of marriage, disability status, Madhesh province whereas associated negatively with males, Janajati ethnic group and no alcohol drink in past month. Care seeking from health care provider is very poor among participants with anxiety and depression. Study indicates increased need to develop intervention and build policies to address higher prevalence of depression and anxiety in Nepal. Implementing support systems and mental health services tailored to the specific needs of the targeted groups and increasing access of population to mental health specialists can play a crucial role in reducing the burden of anxiety and depression in Nepal.

## Data Availability

Datasets are available from https://dhsprogram.com/data/available-datasets.cfm

